# Exploring health inequalities arising from language proficiency; a routine health records study set in England

**DOI:** 10.64898/2026.01.28.26345071

**Authors:** Sarah Yeoh, Mai Stafford

**Affiliations:** Public Health, Brent Council, Brent Civic Centre, London, HA9 0FJ, United Kingdom

**Keywords:** Health inequalities, GP records, hospital admissions, healthcare utilisation

## Abstract

**Background:** Within the UK, more than a million people cannot speak English well or at all. The lack of data on English proficiency means that the link between English proficiency and health status and care utilisation is not comprehensively quantified.

**Objective:** Describe the association between English proficiency and patients’ health status and healthcare utilisation, and demonstrate that GP data can be useful in understanding the health burdens of those with poor language proficiency.

**Methods:** The Northwest London (NWL) Discover-NOW database contains linked, deidentified records from General Practices (GPs), hospitals, and social care in NWL. Using this data, we examined health outcomes and healthcare utilisation of people in Brent who are not proficient in English.

**Results:** Prevalence of age-sex-adjusted cardiometabolic conditions was higher in groups that were not proficient in English or spoke a main language other than English. Primary and secondary healthcare utilisation was also higher in groups that were not proficient in English.

**Conclusion:** This work is the first to quantify healthcare utilisation of those not proficient in English using a large, representative sample in a UK setting. It highlights poorer health outcomes in this group. There is a need to improve provision of language support, starting at registration, which would allow for this group to be better understood.

## 1 Introduction

At the time of Census 2021, 16% of the population of England and Wales had been born outside the UK [1]. Out of this, more than a million could not speak English well or at all [2]. However, English proficiency is regarded as important for integration by both migrants and non-migrants. Migrants with better English abilities reside in less deprived neighbourhoods [3] and have more social support [4]. Migrants who speak English to their children at home are perceived by natives as being more integrated [5].

Language proficiency increases with time spent in the destination country [6, 7] - an effect largely associated with increased use of the language. Such opportunities arise during employment and conversing with others in the community [7]. There is therefore a reinforcing effect where migrants with limited or no English ability are less likely to be employed [6] and more likely to live in language enclaves [3], which in turn limit opportunities for English language use and learning. Children are more likely than adults to learn the language [7] older age is related to lower levels of English proficiency [6, 8]. Women tend to have lower proficiency in the new language compared to men [8, 9].

English proficiency is important for accessing services in the UK, such as healthcare. Research has demonstrated that even those comfortable with English can misunderstand medical terminology [10]. Despite the need for professional language services in the NHS [11, 12], interpreting services are underused, with friends, family, and/or bilingual healthcare professionals filling the gap [13]. [14] notes that healthcare professionals may attempt to act as translators even when their proficiency in the patients’ language is low. In a recent UK study, only 63% of South Asian patients with limited or no English proficiency reported using professional interpreting services in primary care [15]. Interpreter services may also be hard to access; one study found that booking interpreter-assisted appointments required additional language support [16].

Extensive literature from the USA highlights how health outcomes are worse for non-proficient speakers. Low English proficiency has been associated with lower preventative healthcare uptake, such as screenings [17], and vaccinations [18]. While disease prevalence is not necessarily higher in those with low English proficiency, evidence suggests that symptom awareness is poorer [19, 20], resulting in potential under-or misdiagnosis [20]. For non-proficient speakers with diagnosed conditions, disease awareness and management has been found to be poorer [21–26], and health interventions less effective, compared to English-proficient counterparts [27]. However, where patients receive language-concordant care, preventative healthcare uptake [17] and condition management [24, 25] are improved. As to secondary healthcare, a review by [28] concluded that non-proficient patients had fewer specialist physician visits, were more likely to forgo necessary medical care, less likely to receive preventative care, and more likely to be re-admitted into hospital within 30 days.

However, the healthcare system in the USA differs markedly from UK’s universal coverage system. Within the UK, 2011 Census data shows that only 65% of those who could not speak English well or at all had self-reported ‘good’ or ‘very good’ health - compared to 88% of those who could [29]. There is also quantitative evidence that non-English language preference is related to higher rates of chronic heart disease (CHD), but not stroke or hypertension [30], although the latter could be related to underdiagnosis [31]. Evidence on the relationship between English ability and diabetes is mixed [30, 31], as is that between English ability and obesity [30, 32]. However, these studies have focused on the South Asian population [31] and cardiometabolic disease [30–32]. Extant studies have not quantified the relationship between healthcare utilisation and language proficiency. Qualitative studies have focused on healthcare experiences. Those with limited proficiency in English report feeling discriminated due to their poor command of English [33–35]. In some cases, a limited command of English may affect migrants’ understanding of how the NHS works [36], which in turn leads to misuse of the healthcare system such as use of emergency services for non-emergency conditions [37, 38]. How limited English proficiency differentiates migrants’ health and healthcare utilisation from natives in the UK is therefore neither well-described nor well-quantified.

Lack of data on English proficiency has contributed to this evidence gap. Routine healthcare records from primary and secondary care contain some data on English proficiency but have so far been under-used. This study uses healthcare records from patients in Brent, London, an area of high ethnic and language diversity, to quantify levels of health and use of care by language proficiency. A third of residents in Brent speak a main language other than English - the second highest rate of all boroughs in England and Wales - and 7% cannot speak English well or at all [39]. The contributions of this paper are to quantify non-proficient patients’ health status and healthcare utilisation and demonstrate that routine health data can be useful in understanding the health burdens of people not proficient in English.

## 2 Methods

### 2.1 Dataset

The DISCOVER-NOW database contains linked, de-identified records from GPs, hospitals, and social care in Northwest London. Authors did not have access to information that could identify individual participants during or after the study. Apart from health data, the database also contains demographic information, such as birth month, sex, first language, and index of multiple deprivation (IMD) decile - which is a small-area measure of relative deprivation across England and Wales. The data were accessed for research purposes on 1^st^ June 2025. As of June 2025, there were records for 2,872,782 currently registered, living patients in the database.

Since children generally attend appointments with parents, parents’ English proficiency is likely to matter more than children’s English proficiency. However, such parent-child relations are difficult to determine from the routine health records. Thus, children under the age of 18 were excluded from the sample. The final analytical sample was formed by patients over the age of 18, with a known, recorded gender, living in Brent, registered with a GP in the Brent health borough (previously Brent Clinical Commissioning Group), and alive from 1 January 2024 to 31 December 2024.

### 2.2 Measures

#### 2.2.1 Language proficiency

Any patient with any GP record since 2010 of needing a translator, being unable to read or speak English, or speaking English poorly was categorised as not being proficient in English (henceforth, NP, Figure 1). It is noted, however, that language proficiency increases with time spent in the destination country [6, 7]. A previous study found that the probability of self-reported English proficiency was significantly higher starting from the 4th year since migration, compared to the year of migration [7]. Thus, patients who had such a GP record previously may have since improved their English language proficiency. Patients who had such a record 4 or more years ago were therefore separately categorised as being not proficient in English previously (henceforth, NP-Prev).

**Figure 1.**
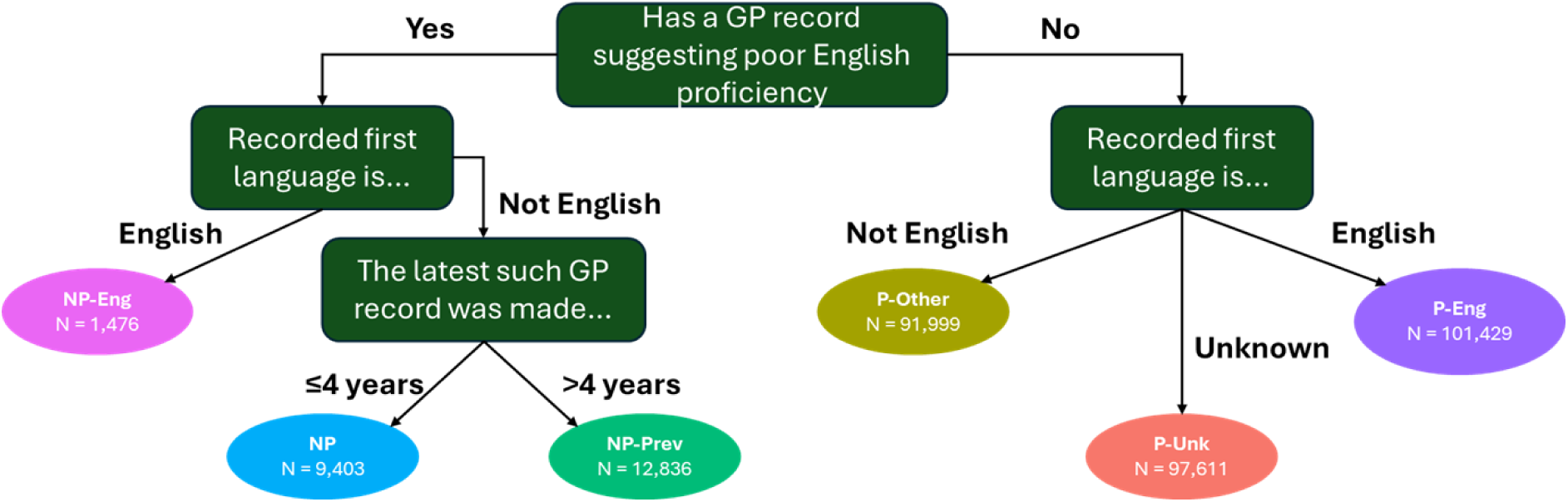
Categorising English proficiency P-Eng = Proficient in English with English as their first language; P-Unk = Proficient in English, unknown first language; P-Other = Proficient in English, but English is not their first language; NP-Prev = Not proficient in English, captured previously in GP record; NP = Not proficient in English; NP-Eng = English as their first language plus a record indicative of poor English proficiency

It is also possible that there are patients who are not proficient in English, but who did not have such a record. Moreover, even if migrants have good levels of English ability, their health and care status may differ from locals. Patients with a first language other than English are likely to be such migrants. As such, patients who did not have any records indicating poor English ability (i.e. the assumed ‘Proficient’ group) were further subdivided on the basis of whether their first language was English (P-English), other than English (P-Other), or unknown (P-Unknown).

There were 1,476 (0.5%) residents who had English as their first language but also had a record indicative of poor English proficiency. This group was designated ‘NP-English’. Of the NP-English group, 1,246 had an entry in their GP records indicating an interpreter was needed.

#### 2.2.2 Health and healthcare utilisation

Discover-NOW also includes de-identified records from GPs on whether an individual has long-term conditions (LTCs) and LTC diagnosis date. LTCs assessed in the primary care quality outcomes framework were included here.

Four measures were used as indicators for healthcare utilisation: number of GP encounters, number of emergency department attendances, number of avoidable emergency department attendances, and number of hospital admissions. Number of GP encounters was proxied by number of distinct days with an entry in the GP record. While this included administrative entries where there were no patient-GP interactions, this over-counting should nonetheless be similar across both proficient and non-proficient groups. Similarly, number of emergency department attendances and number of hospital admissions were proxied by number of distinct start dates. Emergency department attendances with a Healthcare Resource Group (HRG) code indicating no further investigation or significant medication were labelled as an “avoidable attendance”, based on the assumption that the issue could potentially have resolved in primary care. These measures were calculated for the period from 1 January 2024 to 31 December 2024.

### 2.3 Statistical analyses

To compare LTC prevalence, age-and gender-standardised ratios were calculated by indirect standardisation. Age-and gender-specific rates of the P-English group were applied to the age and gender composition of the five other language groups to derive expected counts. These expected counts were then compared against observed counts to derive ratios.

As there was a positive skew in the number of GP encounters, a log-transformation (plus one) was applied. A linear regression model was then built for the transformed model, adjusting also for age, gender, deprivation in the area of residence (based on the Index of Multiple Deprivation in the lower super output area), and number of LTCs.

For number of emergency attendances, avoidable emergency attendances, and hospital admissions, the measures were dichotomised, creating measures indicating whether each patient had had one attendance (admission) or not, and whether they had had multiple attendances (admissions) or not. A separate logistic regression model was created for each of these six binary measures, and also included age, gender, deprivation, and number of LTCs. Additionally, for the avoidable emergency department attendances regression models, patients who had not had any emergency department attendances were excluded from the model sample.

There may be ethnic differences in English language proficiency. Ethnic inequalities in health are also well-documented. As such, a sensitivity analysis was conducted by creating regression models that also adjusted for ethnicity to control for potential confounding, in addition to the above-mentioned variables.

## 3 Results

### 3.1 Demographics

The search yielded 314,754 residents in Brent over the age of 18, of whom 9,403 (3.0%) were NP, 12,836 (4.1%) were NP-Prev, 91,999 (29.2%) were P-Other, 97,611 (31.0%) were P-Unk. In the 2021 Census, 26.2% of the population did not have English as their main language but could speak English well, and 7.5% of the population could not speak English well or at all [40].

Demographics are presented in Table 1. All non-proficient groups had more females (NP: 58% females; NP-Prev: 57% females; NP-English: 54% females) than males. The P-English group had a roughly equal proportion of males and females, while the other proficient groups had more males than females. The P-Unknown group was the youngest (median age 37 years), while the NP-Prev group was the oldest (median age 47 years). A lower proportion of the P-Unknown and P-Other group lived in the most deprived deciles, while almost a quarter of the NP-English group (24.6%) lived in the two most deprived deciles.

**Table 1.**
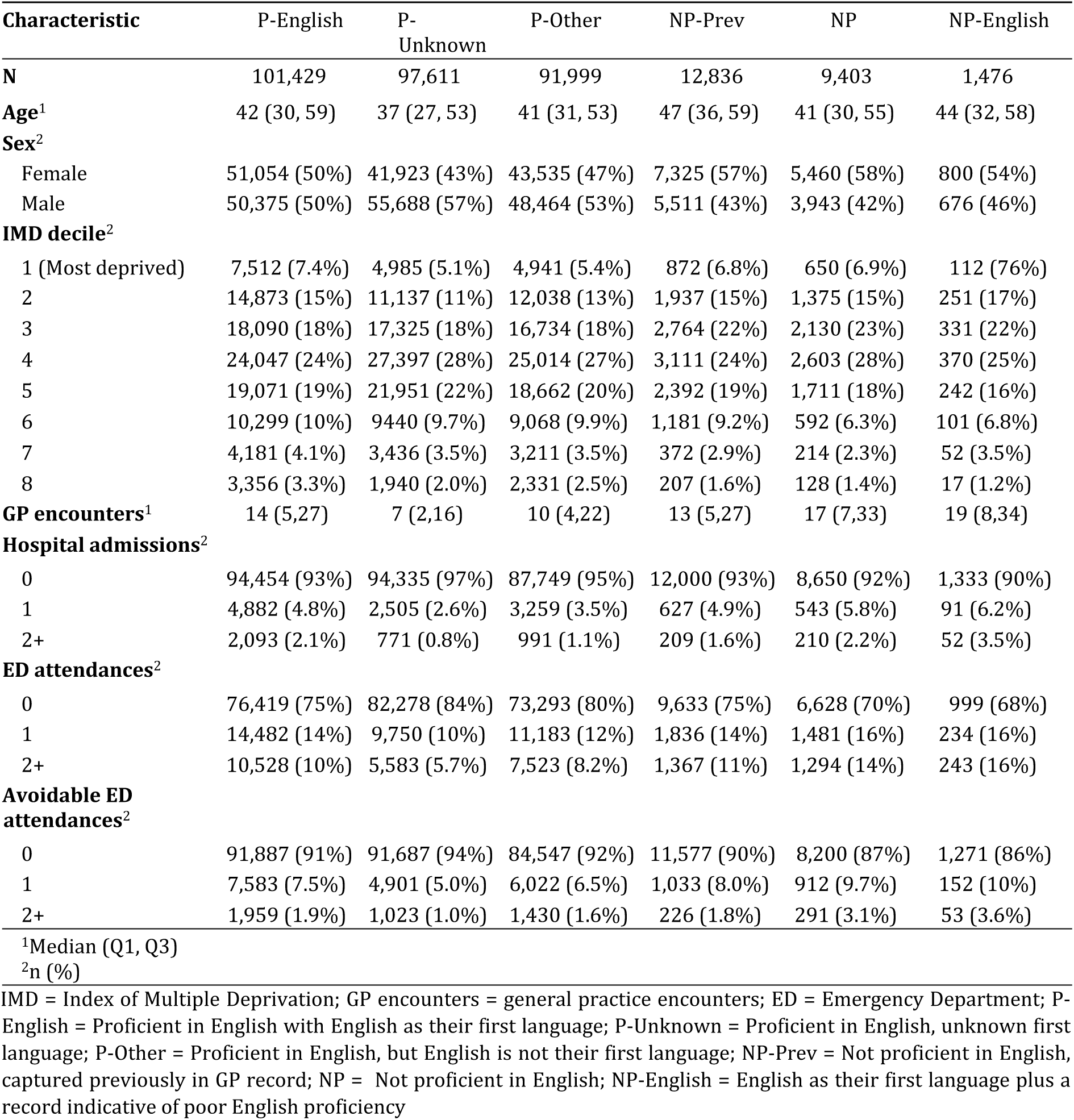
Demographics and healthcare utilisation.

### 3.2 Long-term conditions

Raw (unadjusted) prevalences of long-term conditions are summarised in Table 2. After accounting for age and gender, the P-Other, P-Unknown, and NP-Prev groups had overall lower prevalence of LTC morbidity and multiple long-term conditions compared to the P-English group (Figure 2). Across several LTCs (anxiety, depression, mental health, asthma, COPD, cancer, and CKD), the P-English group had higher prevalence than all other groups less the NP-English group. Outcomes were worst for the NP-English group, which had higher prevalences of various LTCs (including serious mental illness, dementia, epilepsy, and learning disability) compared to the P-English group.

**Figure 2.**
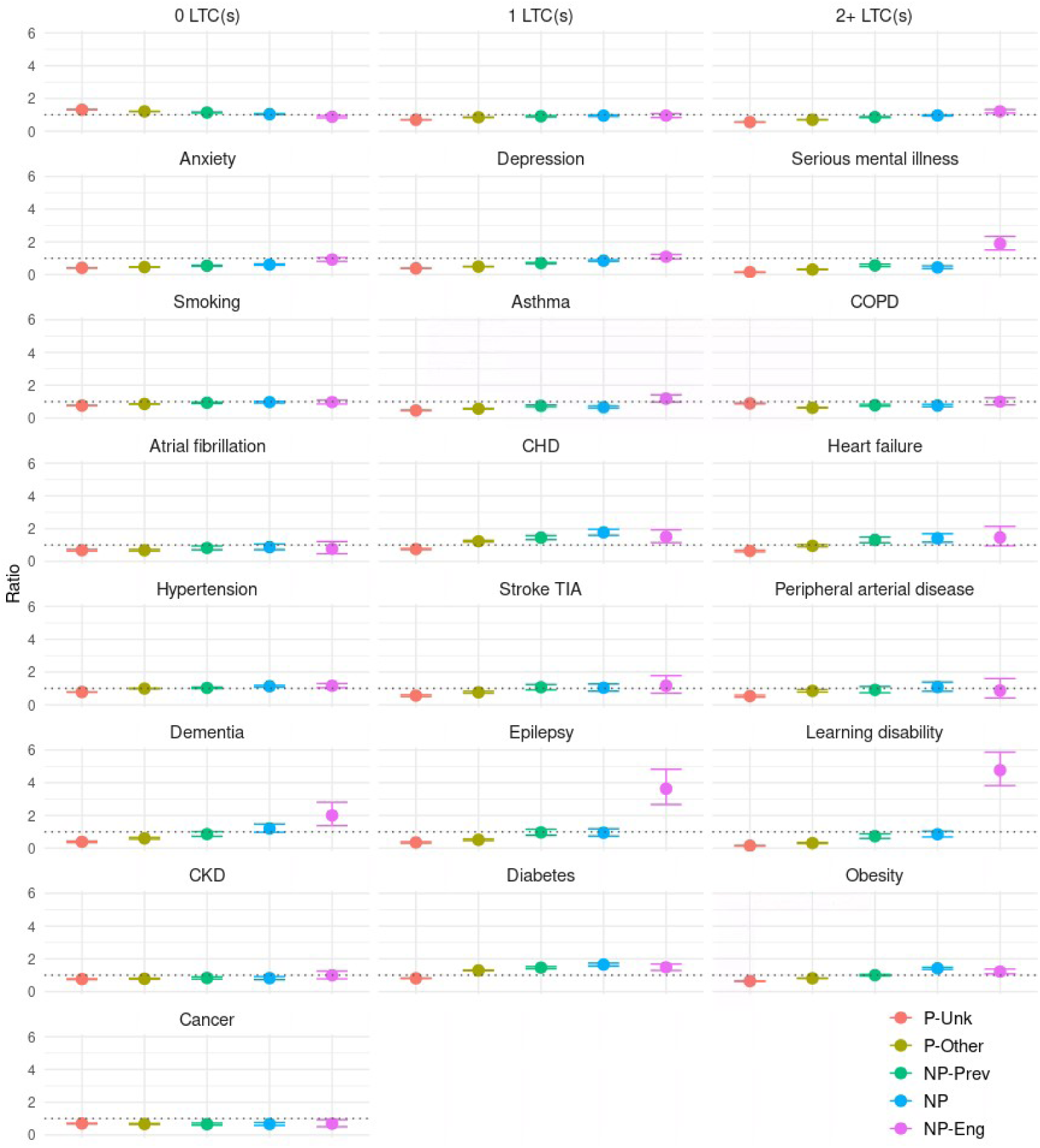
Age-and gender-standardised ratios for prevalence of LTCs by language proficiency P-Unk = Proficient in English, unknown first language; P-Other = Proficient in English, but English is not their first language; NP-Prev = Not proficient in English, captured previously in GP record; NP = Not proficient in English; NP-Eng = English as their first language plus a record indicative of poor English proficiency

**Table 2.** Prevalence of long-term conditions.

| Characteristic<sup>1</sup> | P-English<br>N = 101,429 | P-Unknown<br>N = 97,611 | P-Other<br>N = 91,999 | NP-Prev<br>N = 12,836 | NP<br>N = 8,403 | NP-English<br>N = 1,476 |
| --- | --- | --- | --- | --- | --- | --- |
| Number of LTCs |  |  |  |  |  |  |
| 0 | 50,102 (49%) | 70,563 (72%) | 58,958 (64%) | 6,790 (53%) | 5,018 (53%) | 634 (43%) |
| 1 | 19,324 (19%) | 12,567 (13%) | 14,904 (16%) | 2,340 (18%) | 1,727 (18%) | 274 (19%) |
| 2+ | 32,003 (32%) | 14,481 (15%) | 18,137 (20%) | 3,706 (29%) | 2,658 (28%) | 568 (38%) |
| Anxiety | 17,448 (17%) | 6,577 (6.7%) | 7,227 (7.9%) | 1,251 (9.7%) | 1,043 (11%) | 240 (16%) |
| Asthma | 6,839 (6.7%) | 2,748 (2.8%) | 3,279 (3.6%) | 680 (5.3%) | 413 (4.4%) | 120 (8.1%) |
| Atrial fibrillation | 1,918 (1.9%) | 810 (0.8%) | 768 (0.8%) | 190 (1.5%) | 101 (1.1%) | 19 (1.3%) |
| Cancer | 4,459 (4.4%) | 2,160 (2.2%) | 2,053 (2.2%) | 373 (2.9%) | 219 (2.3%) | 43 (2.9%) |
| CHD | 3,091 (3.0%) | 1,583 (1.6%) | 2,485 (2.7%) | 528 (4.1%) | 356 (3.8%) | 61 (4.1%) |
| CKD | 5,344 (5.3%) | 2,535 (2.6%) | 2,520 (2.7%) | 576 (4.5%) | 288 (3.1%) | 71 (4.8%) |
| COPD | 6,231 (6.1%) | 4,243 (4.3%) | 2,992 (3.3%) | 655 (5.1%) | 383 (4.1%) | 90 (6.1%) |
| Dementia | 1,262 (1.2%) | 291 (0.3%) | 428 (0.5%) | 146 (1.1%) | 94 (1.0%) | 33 (2.2%) |
| Depression | 15,619 (15%) | 5,407 (5.5%) | 7,010 (7.6%) | 1,485 (12%) | 1,308 (14%) | 257 (17%) |
| Diabetes | 10,570 (10%) | 6,408 (6.6%) | 10,290 (11%) | 2,076 (16%) | 1,373 (15%) | 225 (15%) |
| Epilepsy | 891 (0.9%) | 281 (0.3%) | 387 (0.4%) | 109 (0.8%) | 73 (0.8%) | 47 (3.2%) |
| Heart failure | 1,361 (1.3%) | 536 (0.5%) | 768 (0.8%) | 225 (1.8%) | 119 (1.3%) | 26 (1.8%) |
| Hypertension | 19,382 (19%) | 10,772 (11%) | 13,777 (15%) | 2,730 (21%) | 1,699 (18%) | 323 (22%) |
| Learning disability | 1,300 (1.3%) | 196 (0.2%) | 365 (0.4%) | 111 (0.9%) | 100 (1.1%) | 89 (6.0%) |
| Serious mental illness | 3,105 (3.1%) | 410 (0.4%) | 884 (1.0%) | 238 (1.9%) | 124 (1.3%) | 87 (5.9%) |
| Obesity | 14,669 (14%) | 7,860 (8.1%) | 10,334 (11%) | 2,080 (16%) | 1,941 (21%) | 275 (19%) |
| Peripheral arterial disease | 850 (0.8%) | 303 (0.3%) | 473 (0.5%) | 95 (0.7%) | 62 (0.7%) | 10 (0.7%) |
| Stroke or TIA | 1,283 (1.3%) | 471 (0.5%) | 628 (0.7%) | 175 (1.4%) | 90 (1.0%) | 20 (1.4%) |
| Smoking | 20,801 (21%) | 15,812 (16%) | 17,232 (19%) | 2,483 (19%) | 1,842 (20%) | 292 (20%) |

<sup>1</sup>n (%)
LTCs = Long-term conditions; TIA = Transient ischaemic attack; P-English = Proficient in English with English as their first language; P-Unknown = Proficient in English, unknown first language; P-Other = Proficient in English, but English
is not their first language; NP-Prev = Not proficient in English, captured previously in GP record; NP = Not proficient in English; NP-English = English as their first language plus a record indicative of poor English proficiency

All groups, less the P-Unknown group, had higher prevalence of CHD and diabetes, compared to the P-English group. The NP and NP-Prev groups additionally had higher prevalence of heart failure. The NP and NP-English groups also had higher prevalence of hypertension, and obesity. The P-Other group had higher prevalence of CHD and diabetes, while the P-Unknown group had lower prevalence across all LTCs.

### 3.3 Healthcare utilisation

The NP-English group had significantly higher use across all measures (Table 3).

**Table 3.** Association between healthcare utilisation and language proficiency among adults in Brent.

| Characteristic | GP Encounters<br>Beta (95% CI)<sup>1</sup> | Hospital Admissions |  |
| --- | --- | --- | --- |
|  |  | Any<br>OR (95% CI)<sup>1</sup> | Multiple<br>OR (95% CI)<sup>1</sup> |
| **Age** | 0.00 (0.00 0.01)*** | 1.02 (1.02 1.02)*** | 1.03 (1.03 1.03)*** |
| **Sex** |  |  |  |
| Female | - | - | - |
| Male | -0.19 (-0.19, -0.19)*** | 0.93 (0.90, 0.96)*** | 1.07 (1.01, 1.14)* |
| **IMD decile** |  |  |  |
| 1 (least deprived) | - | - | - |
| 2 | -0.01 (-0.02, -0.01)*** | 0.90 (0.84, 0.97)** | 0.90 (0.80, 1.03) |
| 3 | 0.00 (-0.01, 0.01) | 0.82 (0.77, 0.88)*** | 0.76 (0.67, 0.86)*** |
| 4 | 0.00 (0.00, 0.01) | 0.84 (0.79, 0.90)*** | 0.77 (0.69, 0.87)*** |
| 5 | -0.02 (-0.02, -0.01)*** | 0.78 (0.73, 0.83)*** | 0.74 (0.65, 0.84)*** |
| 6 | -0.03 (-0.04, -0.03)*** | 0.74 (0.69, 0.80)*** | 0.63 (0.55, 0.73)*** |
| 7 | -0.02 (-0.02, -0.01)*** | 0.75 (0.67, 0.83)*** | 0.64 (0.53, 0.78)*** |
| 8 | 0.00 (-0.01, 0.01) | 0.88 (0.79, 0.98)* | 0.76 (0.62, 0.93)** |
| **Number of LTCs** |  |  |  |
| 0 | - | - | - |
| 1 | 0.32 (0.32, 0.32)*** | 2.02 (1.92, 2.13)*** | 2.59 (2.32, 2.88)*** |
| 2+ | 0.51 (0.51, 0.52)*** | 4.03 (3.85, 4.21)*** | 6.02 (5.48, 6.62)*** |
| **English proficiency** |  |  |  |
| P-English | - | - | - |
| P-Unknown | -0.10 (-0.10, -0.09)*** | 0.69 (0.66, 0.72)*** | 0.61 (0.56, 0.66)*** |
| P-Other | -0.02 (-0.03, -0.02)*** | 0.84 (0.81, 0.88)*** | 0.71 (0.66, 0.77)*** |
| NP-Prev | -0.03 (-0.04, -0.03)*** | 0.94 (0.87, 1.01) | 0.79 (0.68, 0.91)** |
| NP | 0.12 (0.11, 0.13)*** | 1.30 (1.20, 1.41)*** | 1.24 (1.07, 1.43)** |
| NP-English | 0.07 (0.05, 0.09)*** | 1.35 (1.12, 1.61)** | 1.60 (1.19, 2.10)** |

<sup>1</sup>p<0.05; \*\*p<0.01; \*\*\*p<0.001 Abbreviations: CI = Confidence Interval; OR = Odds Ratio

**Table 3 continued** Association between healthcare utilisation and language proficiency among adults in Brent.
| Characteristic | ED attendances |  | Avoidable ED attendances |  |
| --- | --- | --- | --- | --- |
|  | Any | Multiple | Any | Multiple |
|  | OR (95% CI) | OR (95% CI) | OR (95% CI) | OR (95% CI) |
| **Age** | 1.00 (1.00 1.00)*** | 1.00 (1.00 1.00) | 1.00 (1.00 1.00)*** | 1.00 (1.00 1.00) |
| **Sex** |  |  |  |  |
| Female | - | - | - | - |
| Male | 0.81 (0.80, 0.82)*** | 0.82 (0.80, 0.84)*** | 0.81 (0.80, 0.82)*** | 0.82 (0.80, 0.84)*** |
| **IMD decile** |  |  |  |  |
| 1 | - | - | - | - |
| 2 | 0.85 (0.82, 0.88)*** | 0.82 (0.78, 0.86)*** | 0.85 (0.82, 0.88)*** | 0.82 (0.78, 0.86)*** |
| 3 | 0.76 (0.73, 0.79)*** | 0.74 (0.70, 0.78)*** | 0.76 (0.73, 0.79)*** | 0.74 (0.70, 0.78)*** |
| 4 | 0.68 (0.66, 0.71)*** | 0.64 (0.61, 0.67)*** | 0.68 (0.66, 0.71)*** | 0.64 (0.61, 0.67)*** |
| 5 | 0.67 (0.64, 0.70)*** | 0.60 (0.57, 0.63)*** | 0.67 (0.64, 0.70)*** | 0.60 (0.57, 0.63)*** |
| 6 | 0.67 (0.64, 0.70)*** | 0.59 (0.55, 0.62)*** | 0.67 (0.64, 0.70)*** | 0.59 (0.55, 0.62)*** |
| 7 | 0.66 (0.62, 0.69)*** | 0.59 (0.55, 0.65)*** | 0.66 (0.62, 0.69)*** | 0.59 (0.55, 0.65)*** |
| 8 | 0.65 (0.61, 0.69)*** | 0.56 (0.51, 0.61)*** | 0.65 (0.61, 0.69)*** | 0.56 (0.51, 0.61)*** |
| **Number of LTCs** |  |  |  |  |
| 0 | - | - | - | - |
| 1 | 1.83 (1.79, 1.88)*** | 1.95 (1.88, 2.02)*** | 1.83 (1.79, 1.88)*** | 1.95 (1.88, 2.02)*** |
| 2+ | 2.79 (2.72, 2.86)*** | 3.29 (3.18, 3.40)*** | 2.79 (2.72, 2.86)*** | 3.29 (3.18, 3.40)*** |
| **English proficiency** |  |  |  |  |
| P-English | - | - | - | - |
| P-Unknown | 0.72 (0.70, 0.74)*** | 0.70 (0.68, 0.73)*** | 0.72 (0.70, 0.74)*** | 0.70 (0.68, 0.73)*** |
| P-Other | 0.91 (0.89, 0.93)*** | 0.93 (0.90, 0.96)*** | 0.91 (0.89, 0.93)*** | 0.93 (0.90, 0.96)*** |
| NP-Prev | 1.04 (0.99, 1.08) | 1.05 (0.99, 1.11) | 1.04 (0.99, 1.08) | 1.05 (0.99, 1.11) |
| NP | 1.32 (1.25, 1.38)*** | 1.42 (1.34, 1.52)*** | 1.32 (1.25, 1.38)*** | 1.42 (1.34, 1.52)*** |
| NP-English | 1.36 (1.21, 1.52)*** | 1.56 (1.35, 1.80)*** | 1.36 (1.21, 1.52)*** | 1.56 (1.35, 1.80)*** |

<sup>1</sup>p<0.05; \*\*p<0.01; \*\*\*p<0.001 Abbreviations: CI = Confidence Interval; OR = Odds Ratio
IMD = Index of Multiple Deprivation; LTCs = Long-term conditions; P-English = Proficient in English with English as their first language; P-Unknown = Proficient in English, unknown first language; P-Other = Proficient in English, but English is not their first language; NP-Prev = Not proficient in English, captured previously in GP record; NP = Not proficient in English; NP-English = English as their first language plus a record indicative of poor English proficiency

The NP group had significantly more GP encounters (*β* = 0.12; 95% CI: 0.11, 0.13; *t* = 29.97; *p <.*001), and had higher odds of healthcare utilisation across all measures (Any ED attendance OR: 1.32, 95% CI:1.25 - 1.38; Multiple ED attendances OR: 1.43, 95% CI: 1.34 - 1.52, Any avoidable ED attendance OR: 1.20, 95% CI: 1.11 - 1.30; Multiple avoidable ED attendances OR: 1.36, 95% CI: 1.19 - 1.54; Any admission OR: 1.30, 95% CI: 1.20 - 1.41; Multiple admissions OR: 1.24, 95% CI: 1.07 - 1.43).

The NP-Prev group had significantly fewer GP encounters (*β* =-0.03; 95% CI: 0.04, - 0.03; *t* =-9.90; *p <.*001), and were more likely to have an avoidable ED attendance (OR: 1.09, 95% CI: 1.01 - 1.18). They were, however, less likely to have multiple hospital admissions (OR: 0.85, 95% CI: 0.74 - 0.97) (Table 3).

After adjusting for age, sex, deprivation, and number of LTCs, the P-Unknown group had lower healthcare use across all healthcare utilisation measures, while the P-Other group also had significantly lower healthcare use across all measures less multiple avoidable ED attendances (Table 3).

In models that were additionally adjusted for ethnicity, trends were similar, less that the P-Other group did not have significantly higher odds of having had an avoidable emergency department attendances, the NP-Prev group had significantly higher odds of having had any (OR: 1.08, 95% CI: 1.04 - 1.13) or multiple emergency department attendances (OR: 1.10, 95% CI: 1.03 - 1.17), and the NP-English group did not have significantly higher odds of having had an avoidable ED attendance. Results are included in Supplementary material S2 Table.

## 4 Discussion

Based on diagnoses from GP records, the prevalence of LTCs and multiple LTCs was higher in the P-English group than all other comparison groups, less the NP-English group. Poorest health status was seen for the NP-English group, which had higher prevalence for serious mental illness, CHD, hypertension, epilepsy, dementia, diabetes, obesity, and learning disability, compared to the P-English group. After adjusting for age, sex, deprivation, and number of LTCs, the NP-English group also had higher GP and emergency department utilisation, and higher rates of hospital admission.

For the other two non-proficient groups (i.e. NP and NP-Prev), the prevalence of LTCs and multiple LTCs was generally lower. The prevalence for various LTCs (e.g. anxiety, depression, mental health, asthma, and cancer) was lower in the non-proficient groups compared to the P-English group. However, prevalence of cardiometabolic conditions tended to be higher for the non-proficient groups. Controlling for age, sex, deprivation, and LTCs, the use of healthcare services was higher across all measures for the NP group. For the NP-Prev group, usage was mixed, with fewer GP encounters and lower odds of multiple hospital admissions, but higher odds of having had an avoidable emergency department attendance.

The P-Other group corresponds most closely to the non-English language preference group described in [30]. Similarly to [30], this group had higher rates of CHD compared to the P-English group (or the English language preference group), and no difference in rates of hypertension. However, where [30] found higher rates of obesity and no difference in rates of stroke, rates of obesity and stroke are lower in the P-Other group here. The P-Other group also had fewer GP encounters, and lower odds of any or multiple emergency department attendances and any or multiple hospital admissions.

The NP-English group may have the lowest level of English proficiency of all groups. Members of this group were recorded as having English as their main language. However, more than 80% of this group had a record indicating an interpreter was needed - implying poor or no English ability. This contradiction could perhaps be explained by the fact that first language is typically recorded at registration and language barriers arising at the point of GP registration - which is often in English (e.g. [41, 42]) - can lead to inaccurate recording of language preference. Individual language barriers can be ameliorated with the aid of friends and/or family [13]. Yet, the inaccurate recording of language preference at the point of registration suggests such aid was not available to members of the NP-English group, highlighting that the NP-English group may therefore reflect the group with the lowest English ability and less support with this.

### 4.1 Implications

There is a need to improve uptake of interpreting services by signposting to them, providing easy access, more proactively offering interpreting services, and improving their quality. [15] notes that interpreting services continue to be underused compared to the need for them. The NP-English group additionally reflects that there is a clear and urgent need for interpreting services for some even when English is the main language, given their poor health status and high use of healthcare. Apart from this group, other non-proficient groups would also benefit from the use of interpreting services, since language barriers have been reported to negatively affect primary healthcare experience [33–35] and our study highlights higher use of secondary healthcare in the NP group.

This analysis has highlighted cardiometabolic conditions among those with low proficiency in the local setting. This shows where signposting, provision and training of interpreter services could be most beneficial locally. This analysis could be replicated using similar routine healthcare data in other localities to inform public health, general practice, other community and hospital teams about where to target interpreter provision and improvement.

While GP data can be a valuable resource for researching and understanding populations, accuracy in data recording should be improved. The scale of GP data means that even at smaller geographies such as local authorities, niche population groups can be studied at scale, which is invaluable to understanding health and planning interventions. However, the NP-English group suggests that, starting at the point of registration, there may be inaccuracies in recording. Indeed, in health records, inaccuracies and inconsistencies in recording tend to occur more frequently in minoritised groups [43]. Improving recording is key to improving our understanding of overlooked groups, and the first step in addressing and tackling health inequalities.

### 4.2 Limitations

A limitation of this study is the likelihood that there are under-estimates where non-proficient English speakers are concerned. This arises in two areas: under-count of non-proficient English speakers, and under-diagnosis of some LTCs. Registered patients with an entry indicating non-proficiency are categorised as non-proficient (Section 2.2.1). Additionally categorising on the basis of recorded first language provides a further filter to proxy whether patients are proficient in English. However, as the NP-English group demonstrates, there are also patients for whom their first language is incorrectly-recorded as English. It is therefore also possible that a registered patient can be a non-proficient English speaker, but have their first language incorrectly-recorded as English, and no record indicating non-proficiency. Together, these suggest that there could be more non-English proficient speakers not captured, and the number of non-proficient English speakers is an under-estimate of the true figure. Secondly, some languages may not have equivalent terms to describe LTCs [44]. For instance, Urdu does not have an equivalent concept for’anxiety’ [45], while Gujarati has no equivalent concept for’depression’ [46]. Indeed, in languages (e.g. Somali, Bengali) that do not have equivalent conceptualisations of mental health conditions, mental health conditions tend to be conflated as a stigmatised sane-insane dichotomy, which inevitably discourages help-seeking behaviour and diagnosis [45, 47, 48]. These languages are spoken by a large proportion of this study’s sample, and may therefore contribute to under-diagnosis of some LTCs.

### 4.3 Further research

An avenue for future work would be to examine English language proficiency for an even wider set of first languages and ethnic groups. Both this work and [30] investigated language proficiency and health within borough-level settings. When comparing the first languages spoken in this study and [30], the differences in ethnic composition are apparent. European languages (e.g. Portuguese, Spanish, French, Polish) are spoken by more than half the non-English cohort in [30]. However, in this study, South Asian languages and Arabic comprise the most common first languages. Poor English language proficiency is not homogenous, and [7] highlights that greater linguistic distance (e.g. between English and Arabic, vs. between English and French) increases the difficulty and cost of language acquisition. The differences in the most common first languages - especially since neither of these studies has complete direct information on language proficiency - could contribute to some differences in results.

Finally, this study presents a cross-sectional perspective on the differences in health status and healthcare use. GP data provides an opportunity to examine and track anonymised individuals to understand how their health changes over time. Further study could therefore follow these groups - in particular, the P-English, P-Other, and NP groups - to understand how their health status and healthcare use changes over time.

## Data Availability

Data cannot be shared publicly because they were provided to researchers who met the criteria to access confidential data for a specified access period. Data can be applied for by application to the DiscoverNow North West London Data Access Committee. https://www.nwlondonicb.nhs.uk/professionals/digital-and-it/whole-systems-integrated-care-wsic/information-de-identified-users

## 5 Conclusion

This work is the first in the UK to quantify the health utilisation of those not proficient in English, and highlights poorer health outcomes in this group, which in turn, reinforces the need for improved provision of language support. This work also demonstrates that GP data can be used to understand the impacts of language proficiency at scale for research and at small geographies for designing and improving language support locally. Better recording would improve this resource, which would facilitate understanding of overlooked groups.

## Supplementary information

Supplementary file includes **Table S1** Recorded first languages and **Table S2** Association between healthcare utilisation and language proficiency among adults in Brent, adjusted for ethnicity.

## 6 List of abbreviations

COPD: Chronic obstructive pulmonary disease
GP: General practice
HRG: Healthcare Resource Group
IMD: Index of Multiple Deprivation
LSOA: Lower-level super output areas
LTC: Long-term condition
NHS: National Health Service
NP: Not proficient in English
NP-Prev: Not proficient in English, captured previously in GP record
NP-Eng: English as their first language plus a record indicative of poor English proficiency
NWL: Northwest London
ONS: Office for National Statistics
P-English: Proficient in English with English as their first language
P-Other: Proficient in English, but English is not their first language
P-Unknown: Proficient in English, unknown first language
UK: United Kingdom

## Declarations

### 6.1 Ethics approval and consent to participate

The NWL Discover-NOW has ethics and HRA approval for all de-identified research REC Reference 23/WM/0196 IRAS ID 333128. The current study was approved by the Whole Systems Integrated Care Data Access Committee (ID-160).

### 6.2 Consent for publication

Not applicable.

### 6.3 Data availability

Researchers, commissioners and industry partners seeking to access the NWL Discover-NOW data can find more information here: https://discover-now.co.uk/how-to-access-the-data/

### 6.6 Authors’ contributions

Mai Stafford (MS) and Sarah Yeoh (SY) conceptualised the research idea. SY conducted the data analysis, with support from MS. Both authors contributed to the interpretation. SY produced the draft of the paper, and MS critically revised the manuscript. Both authors read and approved the final manuscript.

## 6.7 Acknowledgments

We thank the Northwest London Whole Systems Integrated Care De-identified Data team for their continued support with the data resource, metadata and data access. This work uses data provided by patients and collected by the NHS as part of their care and support.

